# Rates of SARS-CoV-2 transmission between and into California state prisons

**DOI:** 10.1101/2023.08.24.23294583

**Authors:** Preeti Dubey, Christopher M. Hoover, Phoebe Lu, Seth Blumberg, Travis C. Porco, Todd L. Parsons, Lee Worden

**Affiliations:** Francis I. Proctor Foundation, University of California, San Francisco, Calif., USA; Department of Medicine, Division of Hospital Medicine, University of California, San Francisco, Calif., USA; CNRS & Laboratoire de Probabilités, Statistique et Modélisation, Campus Pierre et Marie Curie, Sorbonne Université, Paris, France

## Abstract

Correctional institutions are a crucial hotspot amplifying SARS-CoV-2 spread and disease disparity in the U.S. In the California state prison system, multiple massive outbreaks have been caused by transmission between prisons. Correctional staff are a likely vector for transmission into the prison system from surrounding communities. We used publicly available data to estimate the magnitude of flows to and between California state prisons, estimating rates of transmission from communities to prison staff and residents, among and between residents and staff within facilities, and between staff and residents of distinct facilities in the state’s 34 prisons through March 22, 2021. We use a mechanistic model, the Hawkes process, reflecting the dynamics of SARS-CoV-2 transmission, for joint estimation of transmission rates. Using nested models for hypothesis testing, we compared the results to simplified models (i) without transmission between prisons, and (ii) with no distinction between prison staff and residents. We estimated that transmission between different facilities’ staff is a significant cause of disease spread, and that staff are a vector of transmission between resident populations and outside communities. While increased screening and vaccination of correctional staff may help reduce introductions, large-scale decarceration remains crucially needed as more limited measures are not likely to prevent large-scale disease spread.

## 1 Introduction

Prisons and jails, like other dense congregate settings, have been exceptionally fertile ground for the SARS-CoV-2 virus since its introduction into the United States. Prisons were the sites of 27 outbreaks of over 2,000 COVID-19 cases each in the U.S. in the first year of the pandemic [1], and prison outbreaks continue to arise [2]. Rates of COVID-19 infection have been 2.6 times higher than the community rate in prison residents and 1.6 times higher in prison staff [3]. In the 34 facilities operated by the California Department of Corrections and Rehabilitation (CDCR), 50,575 resident cases and 15,259 staff cases were recorded as of October 9, 2021, and 240 residents and 46 staff died from COVID-19 in that time (staff mortality as of November 2021) [3]. Prisons and jails are an “epidemiological pump” [4] exporting cases to surrounding communities at accelerated rates [4–20]. Disease outbreaks are often especially large in correctional facilities due to bad conditions such as overcrowding [11, 12, 15, 16, 18, 19, 21–28], lack of ventilation and access to healthcare [11, 15, 18, 19, 22, 24, 26, 28, 29], and populations with elevated risk factors for severe disease [8, 9, 11, 14, 16, 18, 19, 22, 24, 26, 28].

In addition to expanding and continuing overall disease spread, correctional institutions amplify racial inequities in disease burden. The rate of incarceration is six times as high for Black people as for white people. Similarly, Latinx and indigenous people are incarcerated at a rate three times that of white people [30]. For this reason, outbreaks in prisons tend to amplify disparities in disease burden [31–41], with all its consequences including the mass disabling impacts of post-acute sequelae of COVID-19 (i.e. long COVID) [42–45], even without accounting for disparities among prison residents. Racial differences in incidence and/or mortality within correctional facilities may further exacerbate health disparity [46]. Additionally, counties with more Latinx and Indigenous people and lower average incomes are associated with higher infection rates in correctional facilities located there [30], indicating another contribution of incarceration to racial disparities in disease burden and early death [47]. Overrepresentation of Black prison staff may mean that COVID exposure among staff also contributes to racial inequity in disease burden, as it does among residents [48].

Both the large size of correctional facility outbreaks and their frequency of occurrence contribute to their impact. Here we investigate the routes by which introductions of SARS-CoV-2 occur. Several outbreaks in prisons have likely been sparked via infected staff [49–53], and by transfers of infected residents between facilities [54–57]. For example, the San Quentin outbreak of June and July 2020, which led to 2241 cases and was caused by a transfer of infected residents from the Correctional Institute for Men (CIM) in Chino, California [58]. In many settings staff have been screened but not widely tested [59] Vaccination rates are low among corrections staff [60]. Prison staff have tended to report higher rates of COVID-19 than the surrounding community [48], though lower than among prison residents, suggesting that they may be a vector of transmission from prison to community.

Previous work [61] identified an effect of countywide COVID-19 case rates on prison cases. Research incorporating both staff and resident cases [62, 63] found an association between community cases and staff cases, and between staff and resident cases, in US federal prisons, and recommended containment among staff to stem introductions of COVID-19 and other diseases into prisons. Reduction of prison populations by decarceration was found to decrease the risk of COVID-19 infection in federal prisons [63, 64].

We use publicly available data to estimate the magnitude of flows to and from California state prisons, estimating rates of transmission from communities to prison staff and residents, among and between residents and staff within facilities, and between staff and residents of distinct facilities. Unlike prior studies, we use a mechanistic model, the Hawkes process, reflecting the dynamics of SARS-CoV-2 transmission for estimation and hypothesis testing.

## 2 Methods

### 2.1 Data

Case counts for prison residents and staff from April 1, 2020 to March 22, 2021 were obtained from the UCLA COVID Behind Bars project, which collected publicly available data from the California Department of Corrections and Rehabilitation (CDCR). These data report a daily cumulative number of resident and staff cases at each of the 34 CDCR facilities. Cumulative case counts for each county were obtained from the California Health and Human Services Open Data Portal. A daily count of community cases in each California county was estimated by subtracting prison resident and staff cases in the county from the cumulative count of community cases. Isotonic regression was then used to estimate nondecreasing series of numbers of county, resident, and staff cases, and daily new cases were then calculated as the first difference of each of those series.

### 2.2 Statistical analysis

We modeled transmission between recorded cases using a Hawkes process model (see Appendix A for details). We used maximum likelihood estimation of the Hawkes process’s parameters to estimate the rates of transmission between and within the resident and staff populations within the facilities and between distinct facilities, and from community to residents and staff of a prison in the prison’s county and across the state.

The model included ten parameters describing intensity of transmission among prison resident, staff, and community populations, listed and described in Table 1. The ten unknown model parameters provide the constants of proportionality determining transmission rates between populations (see Appendix A for details).

**Table 1:**
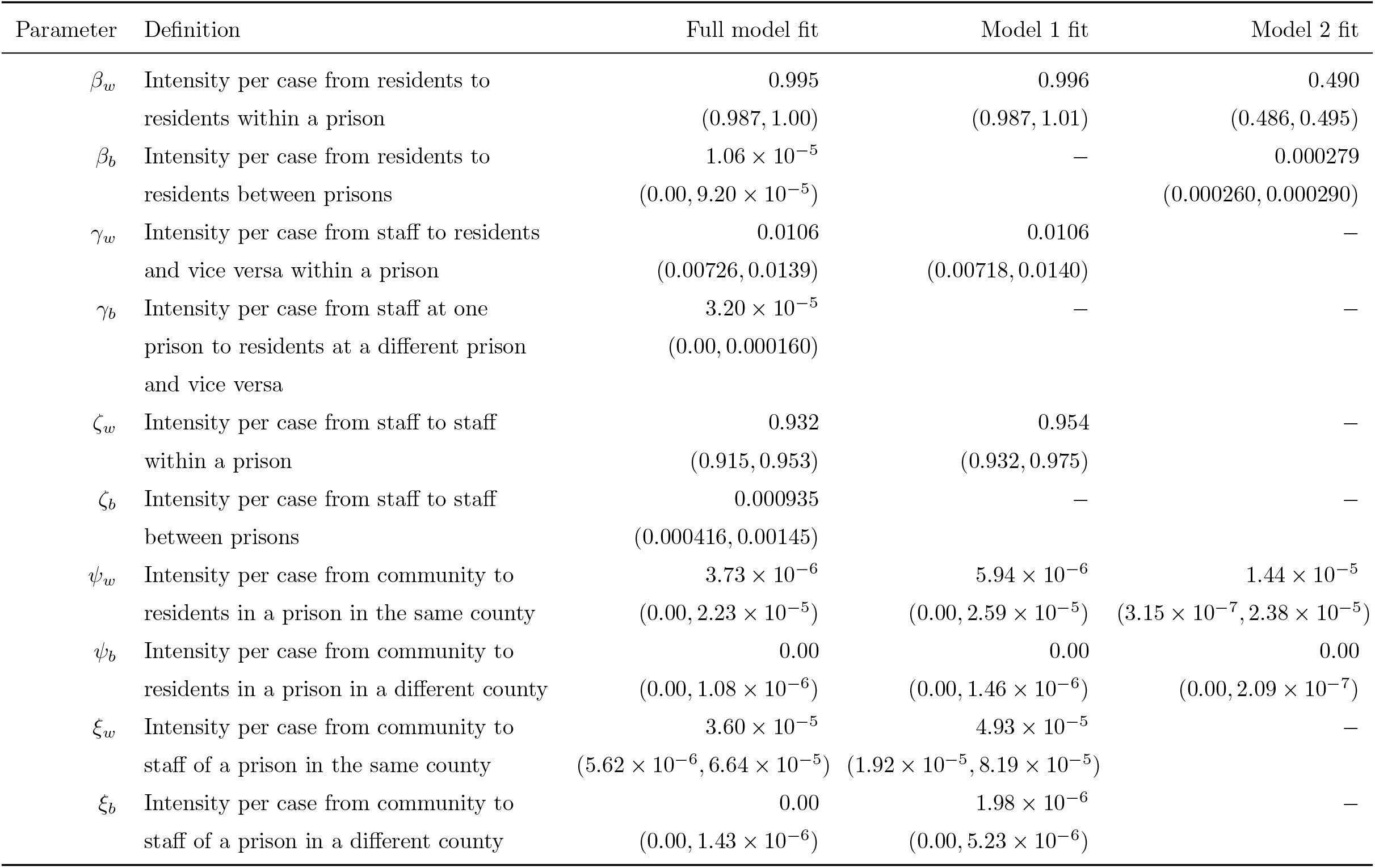
Description and estimated values of model parameters, with 95% confidence intervals.

The timing of transmission events was parametrized using a generation interval between infection of a case and transmission from that case, combined with a reporting interval from a case’s date of infection to the day that case is listed in case count data. The generation time distribution used in these estimates was that estimated in a meta-analysis of COVID-19 generation times [65], a Weibull distribution with mean 5.5 days and standard deviation 1.8 days (parameters *α* = 3.37, *β* = 6.12). The reporting interval was parametrized as a sum of a log-normal incubation period (mean 5.51 days, s.d. 2.4 days) and log-normal detection delay (mean 5 days, s.d. 2.8 days) as estimated by Xu *et al*. [66–68]. The contribution of a source case to creation of secondary cases on each day was assumed proportional to the probability density of the generation interval, plus the secondary case’s reporting interval, minus the first case’s reporting interval. The generation interval and second case’s reporting interval conditional on infection date (*forward reporting interval*) have the above distributions, while the first case’s reporting interval conditional on reporting date (*backward reporting interval*) is estimated as follows, where *p*_*r*_(Δ*t*) is the probability mass function of the forward reporting interval and *Î*(*t*) is an estimate of the daily number of true infections [69]:

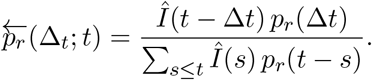

We estimated incidence from the reported case counts as 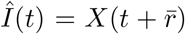 where *X*(*t*) is reported case count on day *t* and 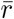 is the nearest whole number to the mean forward reporting delay.

The model parameters were estimated by fitting the data using maximum likelihood estimation, using the log likelihood function defined in Appendix A. Confidence intervals for each parameter value were estimated using profile likelihood estimation [70].

We estimated the number and proportion of resident and staff cases attributed to each of the six different sources of infection modeled—from residents and staff in the same institution, from residents and staff in the CDCR system as a whole, and from community cases in the county containing the institution and across the state. Proportion of cases attributed to a source was estimated by the total intensity of transmission from the source relative to total intensity of transmission from all sources, where total intensity of transmission from a source is the *per capita* transmission rate estimated by the model fit scaled by the total number of cases recorded in the source population.

### 2.3 Transmission hypotheses

To evaluate the implications of the model fit, we constructed two nested models as special cases of the full model by constraining its parameters, detailed in Appendix A.5.

An “independent prisons” model, representing a hypothesis that transmission between prisons is not involved in outbreak dynamics, that is, that outbreaks in prisons originate in introductions from the surrounding communities, into either the resident or staff populations. This model is implemented by defining the between-prison transmission parameters of the full model to equal zero. The remaining parameters were fit using maximum likelihood estimation on the full model’s log likelihood function.

Second, a “well-mixed prisons” model represents a hypothesis that staff and residents do not have distinct roles in transmission, and can be treated as interchangeable in modeling. This is implemented by assuming the contact rates with all populations are equal for staff and for residents.

The remaining free parameters were fit using maximum likelihood estimation on the full model’s log likelihood function.

We applied a likelihood ratio test to evaluate whether there were significant differences in the fit between the full model and either nested model, with three and six degrees of freedom, respectively. We estimated parameter confidence intervals using profile likelihood, and the number and proportion of resident and staff cases attributed to each source, for the two nested models as for the full model as described above.

### 2.4 Sensitivity analysis

Since true dates of infection are unknown and can only be approximated by dates of case detection, there is an inherent amount of uncertainty in the transmission dynamics of the cases recorded that can not be avoided. To examine the dependence of model results on assumptions about precision in detection of cases’ timing, we fit the full model’s parameters using a range of assumed reporting delay distributions, and the best fit results were plotted. The mean and standard deviation of the reporting delay distribution were varied independently by scaling and shifting its probability density function, and the effects of variation in the mean and standard deviation on the model estimate were reported separately.

### 2.5 Code and data availability

All data used in this study came from public sources. Source code and data files are available from the corresponding author by request.

## 3 Results

The data analyzed included 48,389 resident, 17,778 staff, and 2,492,711 community cases after removing prison cases from the California counties’ counts. Outbreaks in California state prisons often coincided with surges of COVID-19 transmission in their surrounding communities (Figure 1), and outbreaks among residents often coincided with surges of cases among staff. The correlations among seven-day average case counts in a facility and its county were 0.42 between residents and staff, 0.24 between community and staff, and 0.14 between community and residents.

**Figure 1:**
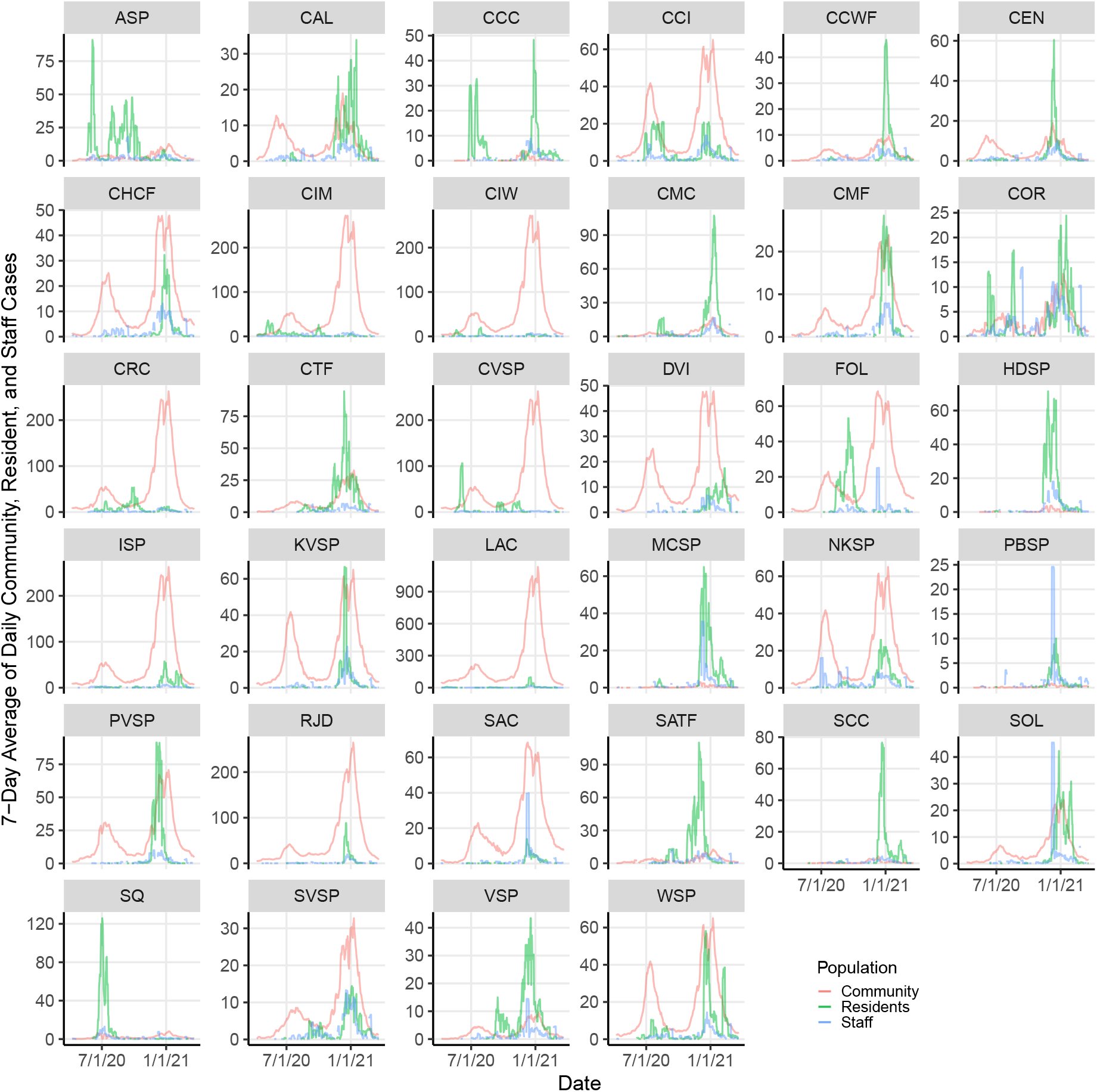
The 7-day average of daily community, resident, and staff cases in each prison and its county, between April 1, 2020 and March 22, 2021. The 7-day average of community cases does not include resident and staff cases in prisons and has been scaled by 1/100. See Table C.1 for abbreviations of facility names.

The full Hawkes process model estimated that transmission to residents was almost entirely from other residents within a facility, and transmission to staff was largely from staff at the same facility (Table 1, Figure 2, Figure 3, Figures B.1–B.6). Infective residents within the same facility are estimated to have contributed about 99.5% of the transmission to resident cases, and similarly staff at the same facility contributed about 92.9% of the transmission to staff. A smaller amount of transmission is estimated between staff across facilities, contributing about 3.1% of transmission to staff. Transmission between residents and staff within a facility is similarly comparatively rare, contributing about 0.4% of resident and 2.9% of staff cases. Contact with the local community contributes about 0.8% to staff cases. The result is similar under the independent prisons model, which assumes no transmission between facilities, and correspondingly attributes more staff cases to local community contacts, while under the well-mixed prisons model, which makes no distinction between staff and residents, transmission to each is primarily from local staff and residents, secondarily from other facilities, and thirdly from local communities.

**Figure 2:**
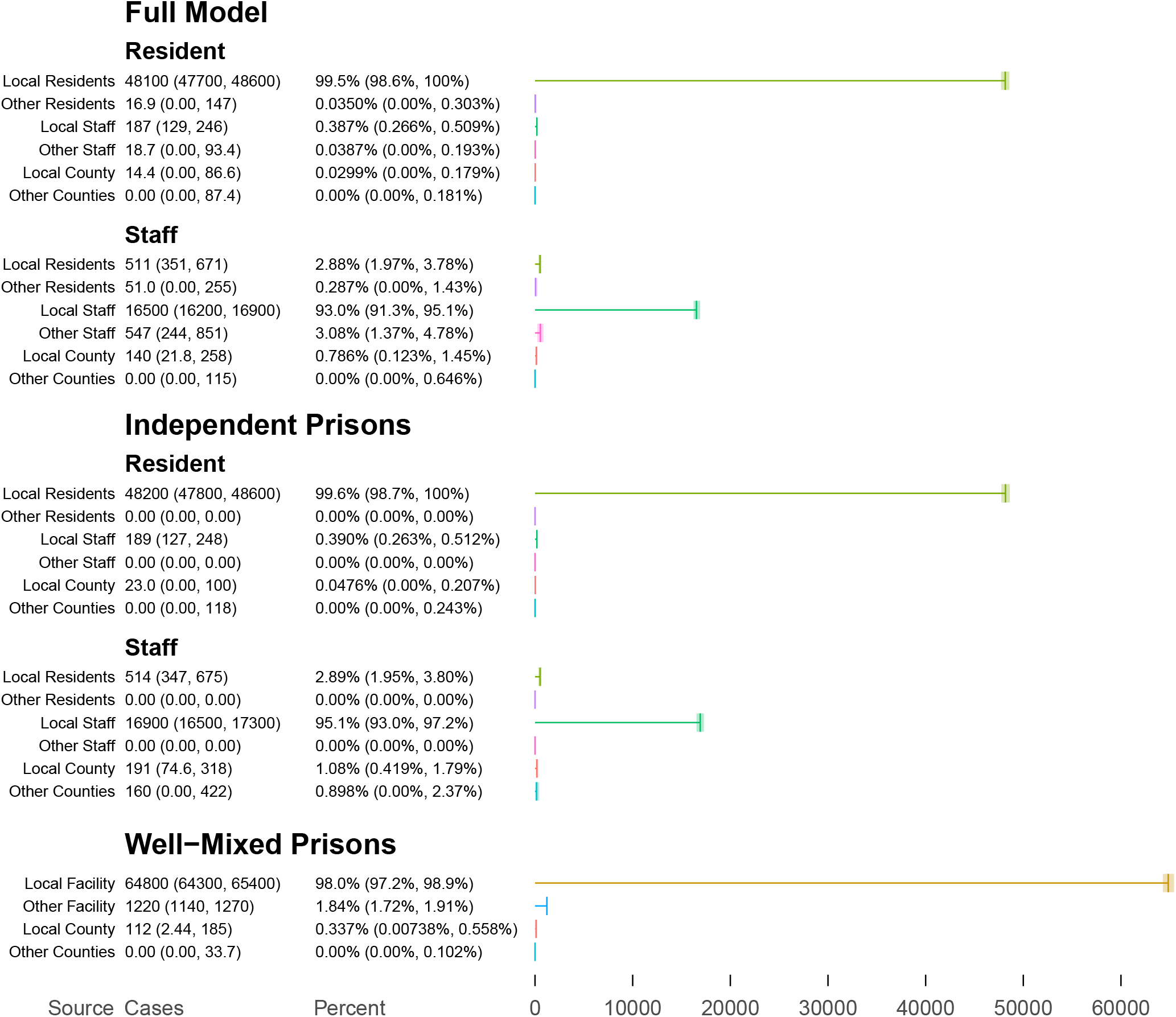
Estimated absolute and percent contributions of sources to transmission to residents and staff cases.

**Figure 3:**
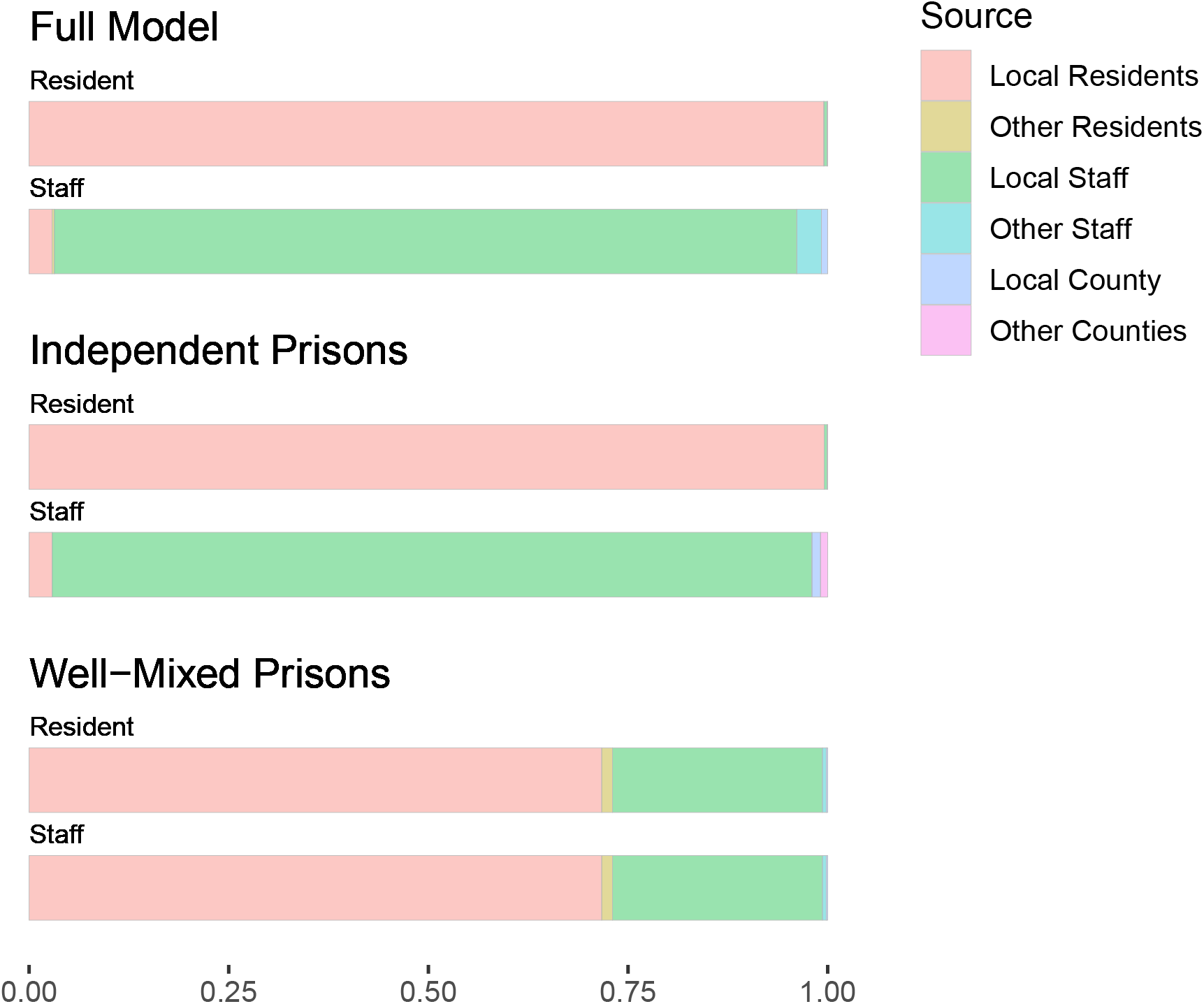
Estimated proportion of resident and staff cases attributed to each source of infection, under the full Hawkes process model and the two nested Hawkes process models.

Likelihood ratio testing found that the full model differed significantly from the independent prisons model (*p <* 0.01), indicating that the rate of transmission between distinct CDCR facilities is significantly different from zero. The rate between staff at distinct facilities in particular has 95% confidence interval distinct from zero in the full model.

The full model also differed significantly from the well-mixed prisons model (*p <* 0.001), indicating a difference between the sources for transmission to staff and to residents. This suggests that transmission from community to staff, and subsequently from staff to residents, is likely a contributor to prison COVID-19 cases.

### 3.1 Sensitivity analysis

We examined how the maximum-likelihood estimate of the full model’s parameters was affected by assumptions about timing of cases’ detection, by varying the mean and standard deviation of the reporting delay distribution. The model results are found to be insensitive to variation in the mean delay. We found that the standard deviation of the reporting delay, indicating the amount of uncertainty in timing of infections, has a quantitative impact on the model results, but not a qualitative one in the range of values examined (Figure 4).

**Figure 4:**
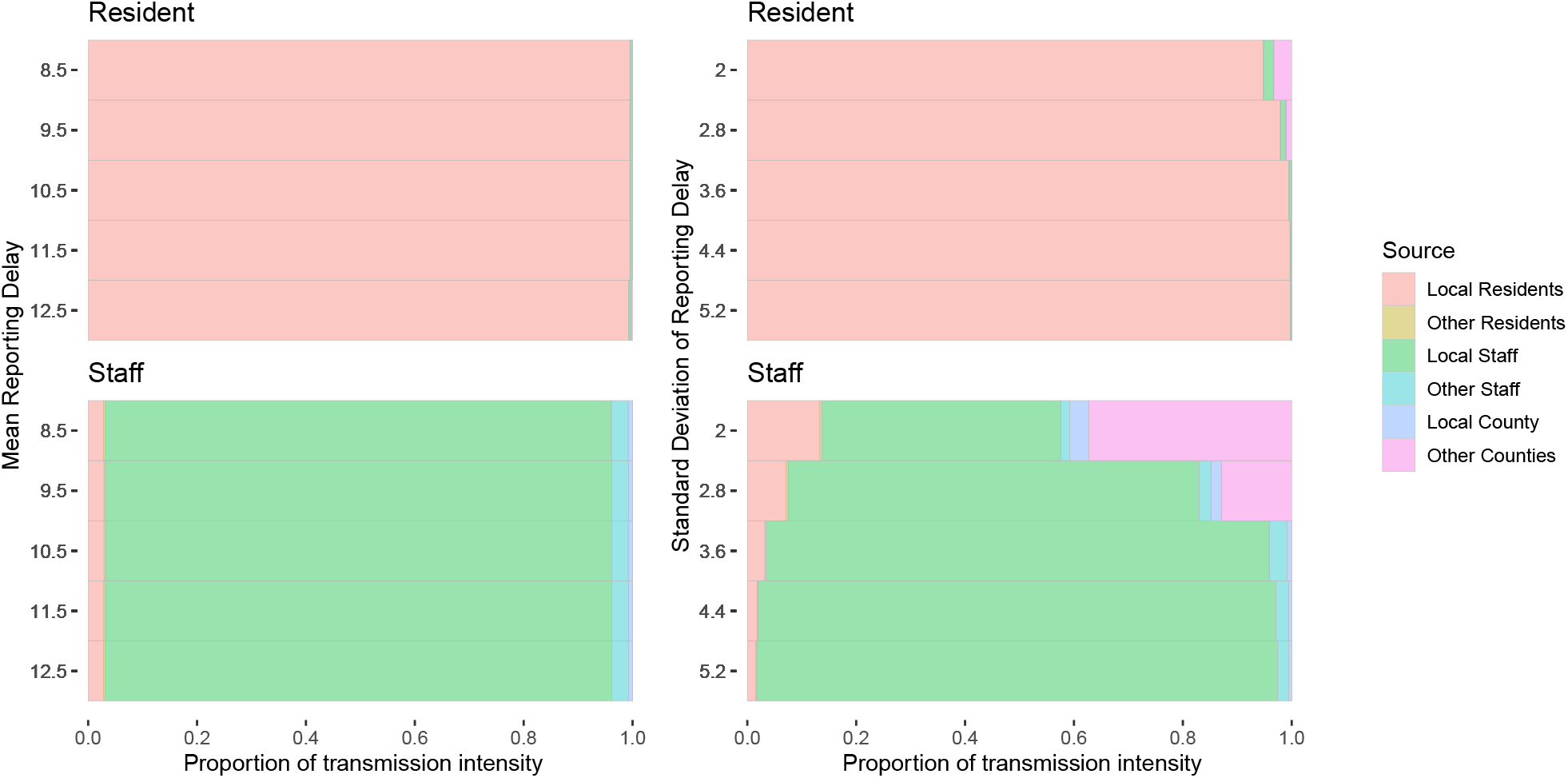
Dependence of full model result on mean (left) and standard deviation (right) of assumed case reporting delay. Tinted bars denote fraction of infection intensity affecting residents and staff due to each source of infection.

## 4 Discussion

Our results indicate that prison staff unsurprisingly have more outside contact events than prison residents, both with the surrounding community and with other facilities. The latter may involve staff who travel between facilities [52, 53]: CDCR data shows staff members work at an average of two facilities [3]. This supports the idea that staff may be an important source of transmission into and out of prisons. Testing of staff may be inadequate to prevent spread into prisons [59], and the problem may be exacerbated by low staff vaccination levels [60].

Both testing and vaccination are important occupational health measures for correctional staff as well as for outbreak prevention [71, 72]. Transmission due to transfer of infected residents is not clearly distinguished from zero by this model, perhaps due to relative rarity compared to everyday transmission. Nonetheless, it is known to have caused major outbreaks in the CDCR system, and must be taken very seriously as a source of risk.

Prevention of prison outbreaks is crucial, not only for protection of residents and staff, but to reduce overall transmission and exacerbation of unjust disparities in disease burden [8, 11, 13, 15, 24, 33, 73–77]. Decarceration continues to be urgently needed [6, 8, 11, 12, 14, 15, 18, 19, 21, 23, 24, 26, 30, 33, 61, 74, 76, 78–97].

This study has a number of limitations. The use of a stationary Hawkes process formulation does not account for temporal variation in transmission rates, whether due to depletion of susceptible individuals or other causes. While we use it here to look at the relative contributions to the transmission rate, which may be relatively insensitive to that approximation, it may introduce inaccuracy. The time period studied may not be predictive of dynamics after March 2021. Differences between the 34 CDCR facilities may be obscured by this model. Dynamics such as rare but influential transfers of infected individuals may not be identifiable from the data set used. Variable case detection could introduce bias; for example, if testing is more frequent during outbreaks in facilities, the proportion of cases originating in the facilities could be overestimated. The assumption of symmetric transmission rates between staff and residents may be a limitation, as for example one or the other population may be more likely to be isolated while infective.

We find the Hawkes process formulation to be a powerful and flexible technique for estimation of transmission rates between segments of a population. Mechanistic knowledge of the process such as a generation time distribution can be used directly to infer mixing rates from data without extensive intermediary steps such as simulation. It can be used to estimate mixing rates in a broad variety of structured-population disease transmission problems, and it may prove useful in a wide range of disease modeling applications in the future.

## Data Availability

All data produced in the present study are available upon reasonable request to the authors

## 4.1 Acknowledgements

PD, TCP, and LW were supported by NIH GM130900. SB, CMH, PL, and TCP were supported by CDC U01CK000590, as part of the Modeling Infectious Diseases in Healthcare Network. SB was also supported by NIH K12 EY031372. LW received partial funding from the Office of the Federal Receiver, which oversees the delivery of healthcare services in California’s state prison system, for research not including this project. The funders had no role in study design, data collection and analysis, decision to publish, or preparation of the manuscript.

### Appendix

#### A Model Details

##### A.1 Hawkes Processes

To estimate the effects of multiple sets of infected cases on one another through time, one can use a Hawkes (or *self-exciting*) process [98–102]. This is equivalent to a branching process, or the assumptions made in *e*.*g*. [103]; Hawkes himself used measles contagion as an example of a self-exciting process.

Hawkes processes assume a intensity function that increases and decreases in proportion to the number of events that have occurred in the past (*e*.*g*. infections are caused by previously infected individuals): if *N* ([*a, b*)) = (# of infection events at times *t* ∈ [*a, b*)) is the empirical measure of the point process, with

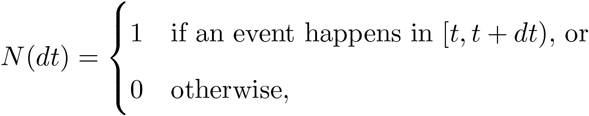

then the intensity function is

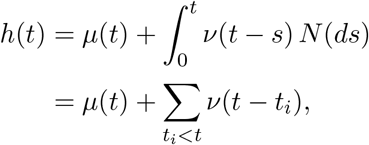

where *t*_*i*_ are the event times in [0, *t*). Here, *μ*(*t*) is an external or deterministic component of the intensity, whereas *ν*(*t*) measures the contribution of past events to the rate of new events. Writing *N* (*t*) = *N* ([0, *t*)) and letting *P* (*t*) be a standard Poisson process, we have

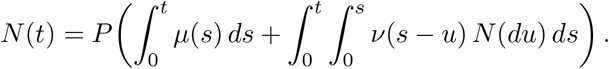

More generally, we can consider mutually exciting processes, *N*_*i*_(*dt*), *i* = 1, …, *m*, with intensities

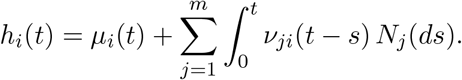

##### A.2 Discrete-Time Hawkes Processes

Implicit in the definition of Hawkes processes is the assumption that one can always resolve the timing of each event, so that no two events are simultaneous. Here, the data consists a series of daily case counts *X*_*i*_(*t*) of each type, and multiple cases are commonly observed each day, so we must define a discrete-time version of the Hawkes process to proceed.

Informally, the Hawkes process is such that Δ*N*_*i*_(*t*) = *N*_*i*_(*t* + Δ*t*) − *N*_*i*_(*t*) is Poisson distributed with rate *h*_*i*_(*t*) Δ*t*, where *h*_*i*_(*t*) depends (only) on events up to time *t*, whereas the probability of multiple events in the interval [*t, t* + Δ*t*) is of order (Δ*t*)^2^, which becomes negligible as Δ*t* → 0. We measure time so that Δ*t* is one day, and thus cannot neglect the possibility of multiple events on a single day, and thus define our process *N*_*i*_(*t*), *t* = 0, 1, … assuming *N*_*i*_(0) is given, whereas Δ*N*_*i*_(*t*) = *N*_*i*_(*t* + 1) − *N*_*i*_(*t*) is Poisson distributed with rate

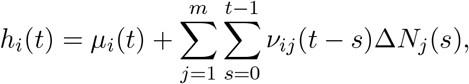

where we can understand the latter terms as the discrete equivalents of the integrals 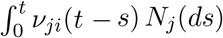. By definition, 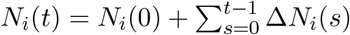.

The probability that Δ*N*_*i*_(*t*) = *n*_*it*_ is then

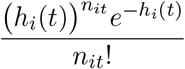

and the log likelihood for a vector of parameters Θ given ***N*** = (*N*_1_, …, *N*_*m*_) is thus

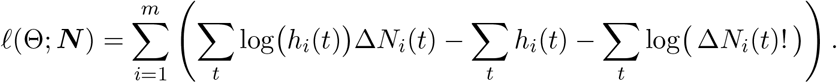

##### A.3 Detection Delays

Inference of transmission dynamics may be affected by the timing of cases’ inclusion in case counts relative to the date of their infection, if the interval from infection to case reporting date is variable.

Let each case, identified by an index *x*, be infected on day *t*_*x*_ and included in case counts for day 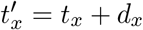, where *d*_*x*_ has probability mass function *p*_*d*_(*s*), *s* = 0, 1, Let *X*_*i*_(*t*) denote the reported case count on day *t* while Δ*N*_*i*_(*t*) = *N*_*i*_(*t* + 1) − *N*_*i*_(*t*) is the number of cases infected on day *t*. The case counts on day *t* are then

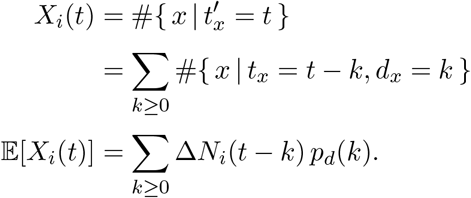

We approximate this process by using a serial interval distribution relating case detections to previous detections. Case detections on day *t* are generated at rate

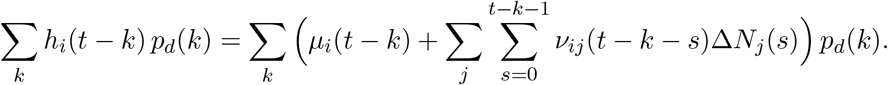

Given a case detected on day *s*, the probability [69, 104] that a given one of its secondary cases is detected on day *t* is

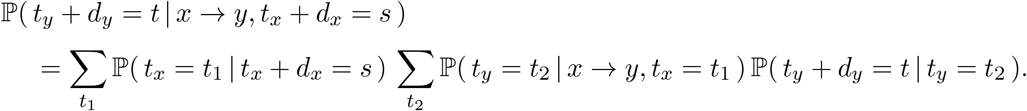

Two of those probabilities are given by distributions already defined:

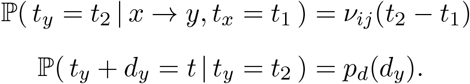

The other one is a *backward detection interval* probability [69]:

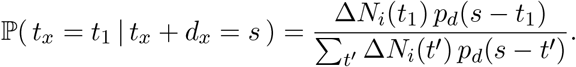

The true daily incidences Δ*N* being unobserved, we approximate by assuming the true incidence curve rises and falls as the reporting curve does with a lag, 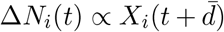, with 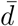 the nearest integer to the mean reporting delay:

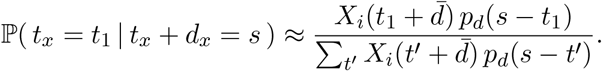

Using that, we can model the cases recorded on day *t* as being generated on day *t* at rate

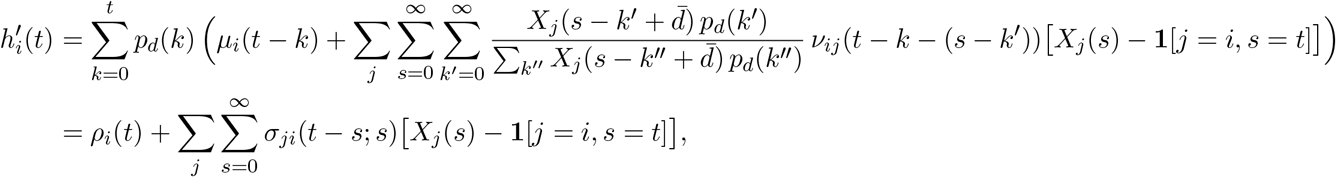

with

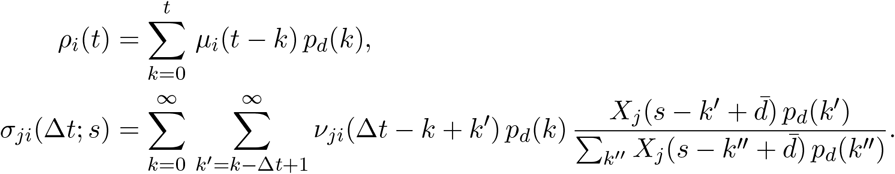

We use this to define an adjusted likelihood function:

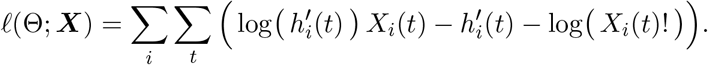

##### A.4 Application to Case Counts

When the process is represented by a series of daily case counts *X*_*i*_(*t*) (*t* = 0, …, *T*) of each type, we have a log likelihood function

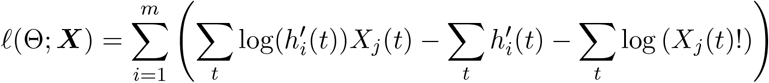

with

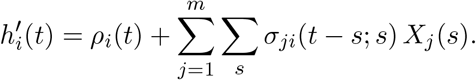

For our purposes, we will assume mutually exciting groups are the number of infected staff at each correctional facility and the number of infected residents at each correctional facility. We will include the number of infected members of each county excluding prison residents and staff as an external source of hazard.

We write 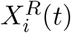 and 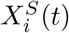 for the daily number of cases reported in residents and staff, respectively, at the *i*^th^ facility, *i* = 1, …, *m*_*f*_ (where *m*_*f*_ is the number of facilities). For the functions *σ*_*ij*_, we use a common generation interval distribution *ν*(Δ*t*) and detection delay distribution *p*_*d*_(Δ*t*) to determine the contribution of each past infection to the present force of infection. Assume that infectious contacts between residents of facility *i* and facility *j* occur at rate *β*_*ij*_ (with units of events per unit time), so that the contribution of the former to infection of the latter on day *t* is 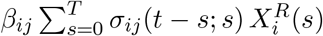. Similarly, assume that infective contacts between staff at facility *I* and residents at facility *j* occur at rate *γ*_*ij*_, such that its contribution to the infection of residents of facility *j* is 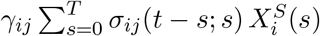. Assume that infective contacts between staff of facility *i* and facility *j* occur at rate *ζ*_*ij*_, so that the contribution to infection of staff of facility *j* due to residents and staff of facility *i* is 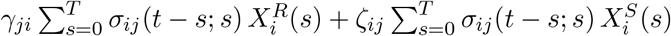.

Finally, we model transmission between the community and the prisons. In this study we consider only transmission into the prisons, not to or between community members, and we assume that all transmission from outside the prison system is from the California county populations. Let *ψ*_*ki*_ denote the contact rate between residents of prison *i* and county *k*, and let *ξ*_*ki*_ denote the contact rate between staff of prison *i* and county *k*. Denote the daily number of (non-prison) cases in county *k* by 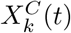, *k* = 1, …, *m*_*c*_, where *m*_*c*_ is the number of counties.

We then write the total intensity for residents in prison *j*, including transmission from residents, staff, and the surrounding community:

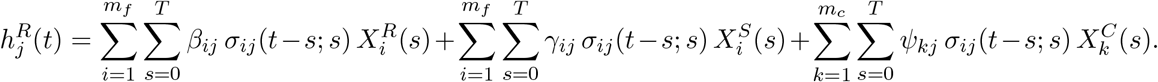

For the intensity for staff in prison *j*, we have

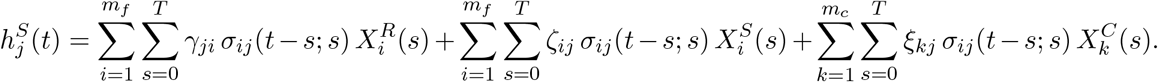

For simplicity, we will assume that *β*_*ij*_ = *β*_*w*_ when *i* = *j* (within facility) and *β*_*ij*_ = *β*_*b*_ when *i* ≠ *j* (between facilities); we use analogous notation for *γ*_*w*_, *γ*_*b*_, *ζ*_*w*_, *ζ*_*b*_. Similarly we model the parameters *ψ* and *ξ* as *ψ*_*w*_ and *ξ*_*w*_ between a facility and the county containing that facility, and *ψ*_*b*_ and *ξ*_*b*_ between a facility and all other counties. Let the parameters be written as a vector Θ = (*β*_*w*_, *β*_*b*_, *γ*_*w*_, *γ*_*b*_, *ζ*_*w*_, *ζ*_*b*_, *ψ*_*w*_, *ψ*_*b*_, *ξ*_*w*_, *ξ*_*b*_). We denote the entire collection of observations 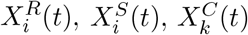 for all *i, k, t* as ***X***. The log likelihood for the parameters is then

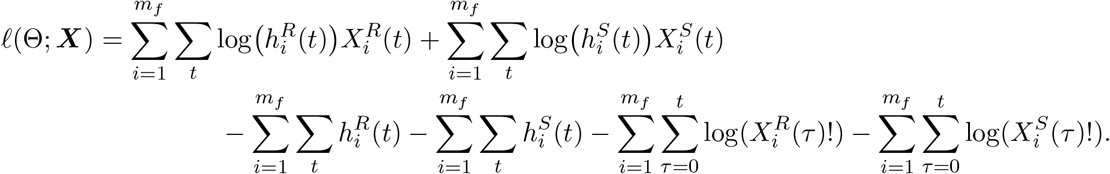

This log likelihood function is used to fit the parameter vector Θ using maximum likelihood estimation.

Given a parameter estimate Θ, the contribution of each source population to transmission to residents or staff is estimated by its contribution to their total Hawkes process intensity over time, 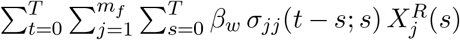 for within-facility transmission from residents to residents. The proportion of transmission from each source is estimated by the absolute contribution divided by the maximum likelihood estimate of the total Hawkes intensity to residents or staff.

##### A.5 Nested models

The independent prisons model was implemented by defining the between-prison parameters *β*_*b*_, *γ*_*b*_, and *ζ*_*b*_ to equal zero. The remaining parameters were used in fitting.

The well-mixed prisons model was implemented by constraining *β*_*w*_ = *γ*_*w*_ = *ζ*_*w*_, *β*_*b*_ = *γ*_*b*_ = *ζ*_*b*_, *ψ*_*w*_ = *ξ*_*w*_, *ψ*_*b*_ = *ξ*_*b*_, and using the remaining free parameters *β*_*w*_, *β*_*b*_, *ψ*_*w*_, *ψ*_*b*_ in fitting.

#### B Detailed transmission intensities

Figures B.1–B.6 detail the estimated intensities of transmission to prison residents and staff attributes to various sources, under the full and nested models described above.

#### C Abbreviations for CDCR facilities

Table C.1 lists the names and abbreviations used for CDCR’s facilities.

**Figure B.1:**
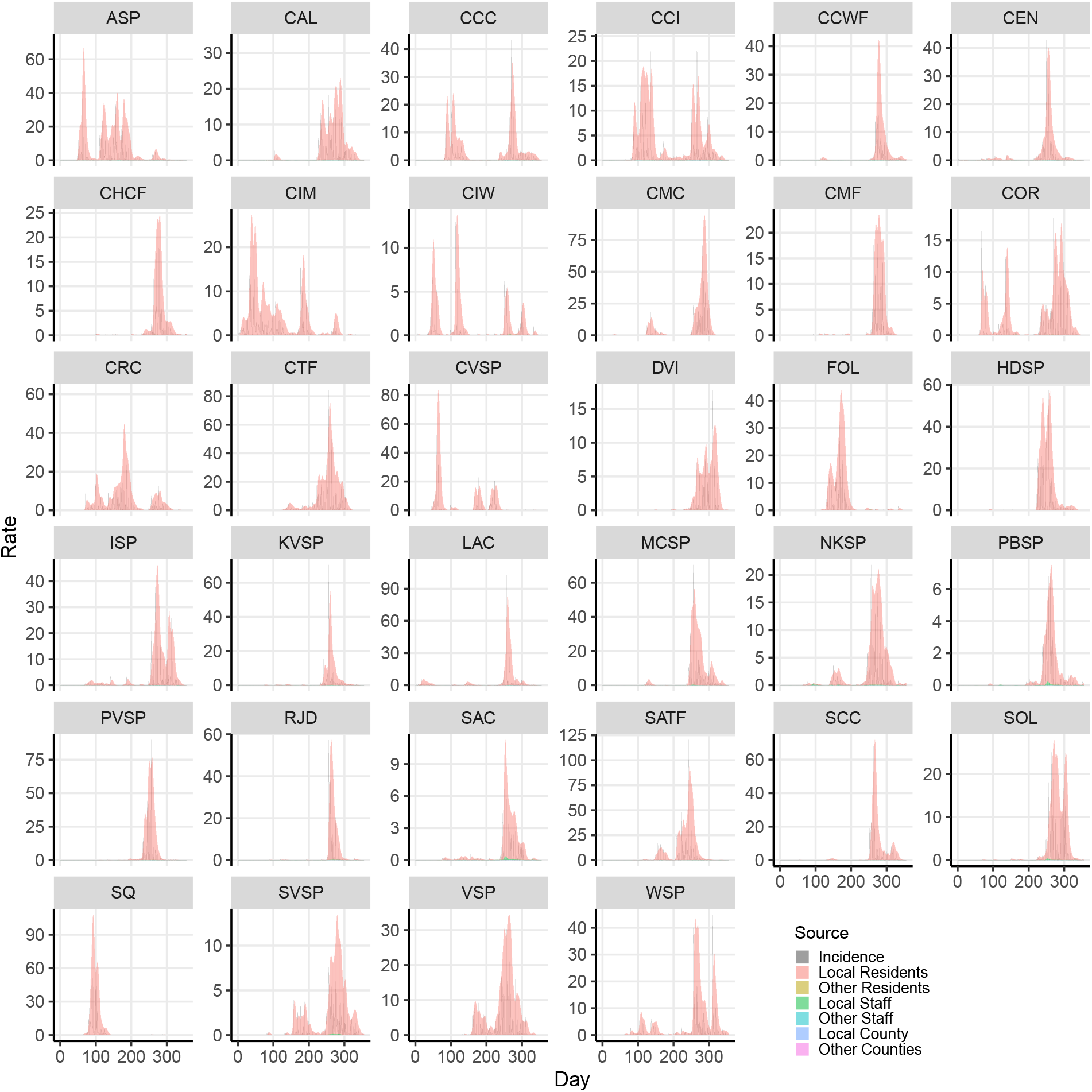
Incidence rates (gray) and intensities (colors, stacked) for resident cases by facility, in the maximum likelihood estimate for the full Hawkes process model.

**Figure B.2:**
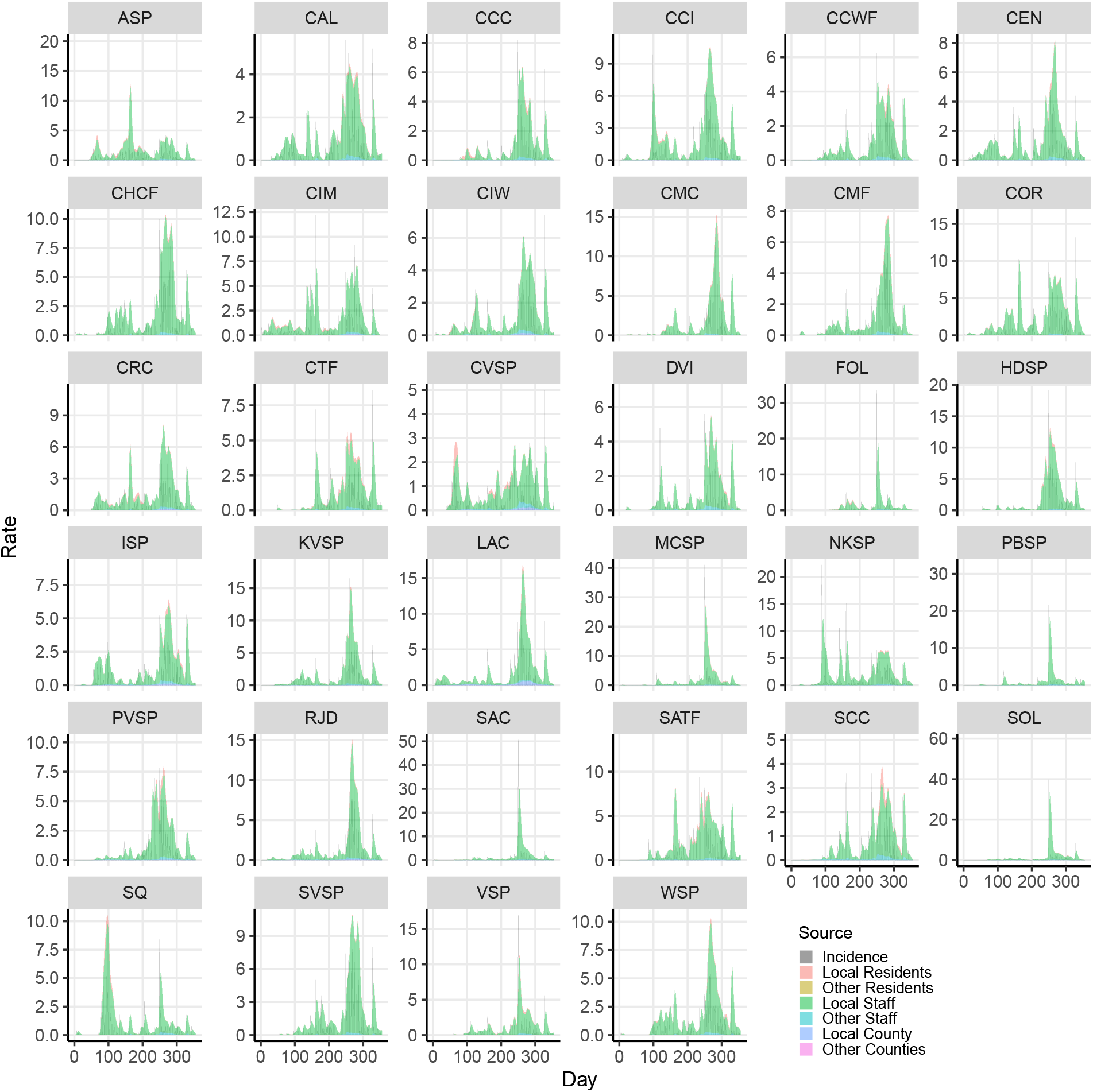
Incidence rates (gray) and intensities (colors, stacked) for staff cases by facility, in the maximum likelihood estimate for the full Hawkes process model.

**Figure B.3:**
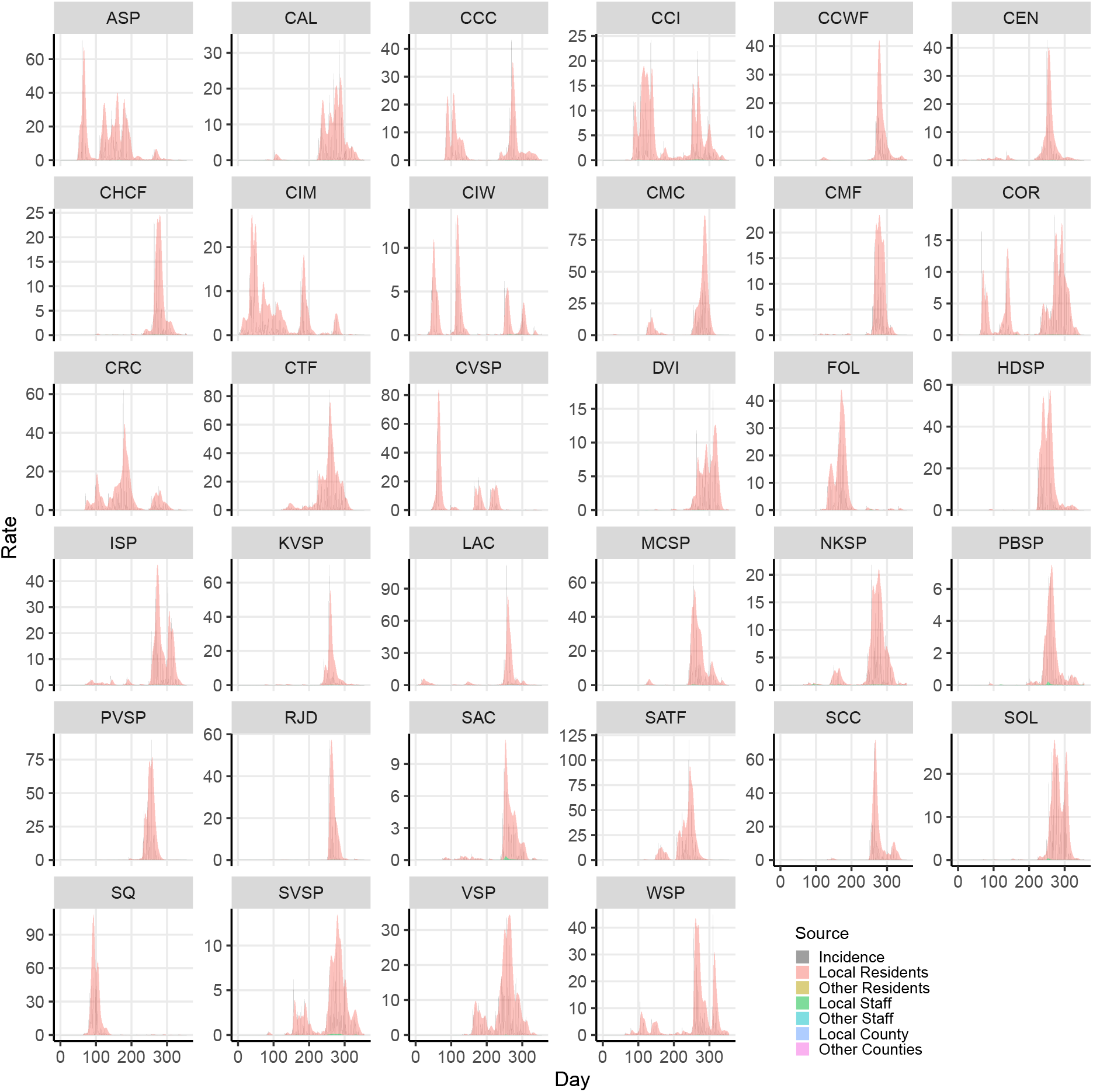
Incidence rates (gray) and intensities (colors, stacked) for resident cases by facility, in the maximum likelihood estimate for the independent prisons nested model, with local transmission only.

**Figure B.4:**
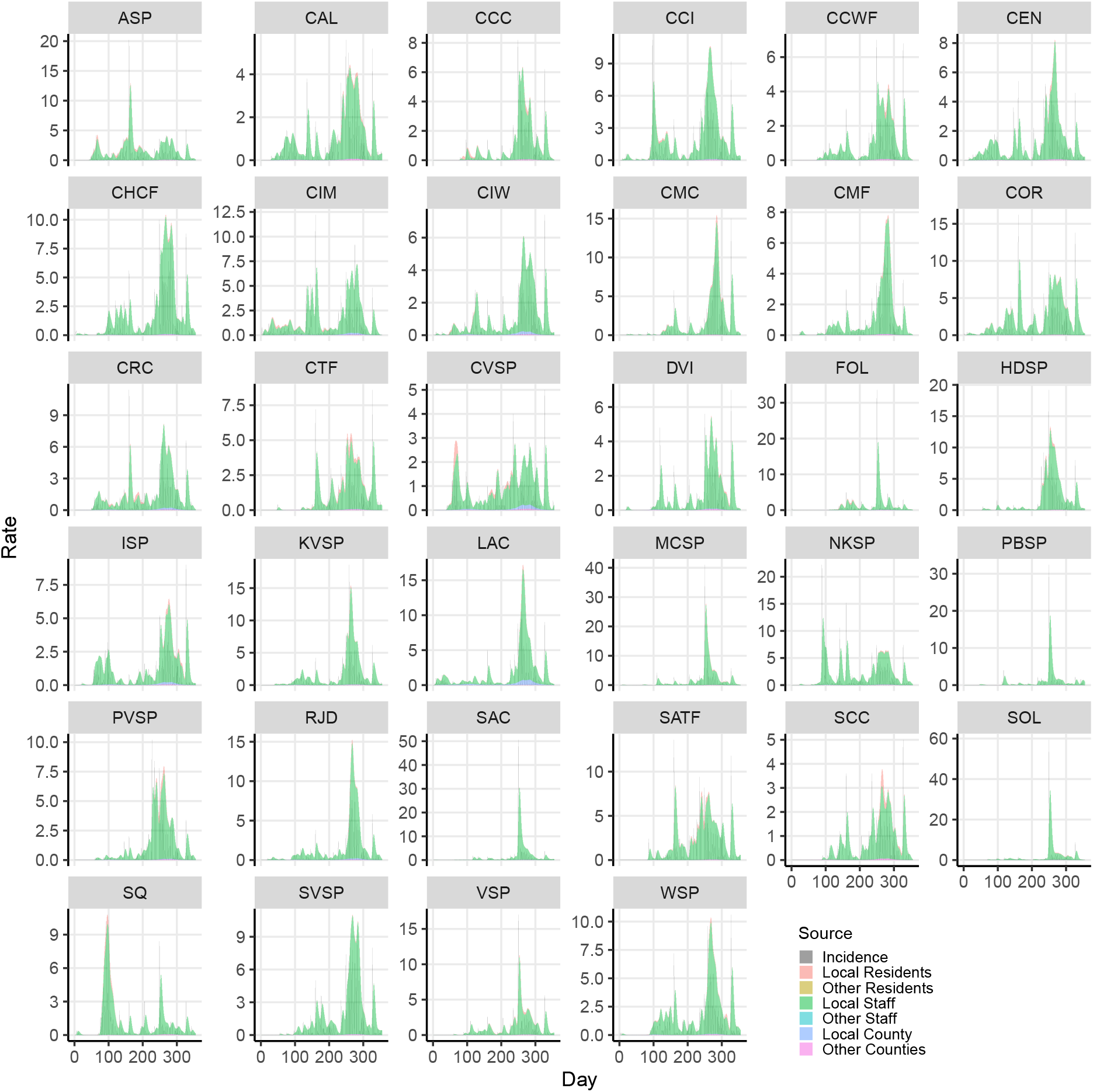
Incidence rates (gray) and intensities (colors, stacked) for staff cases by facility, in the maximum likelihood estimate for the independent prisons nested model, with local transmission only.

**Figure B.5:**
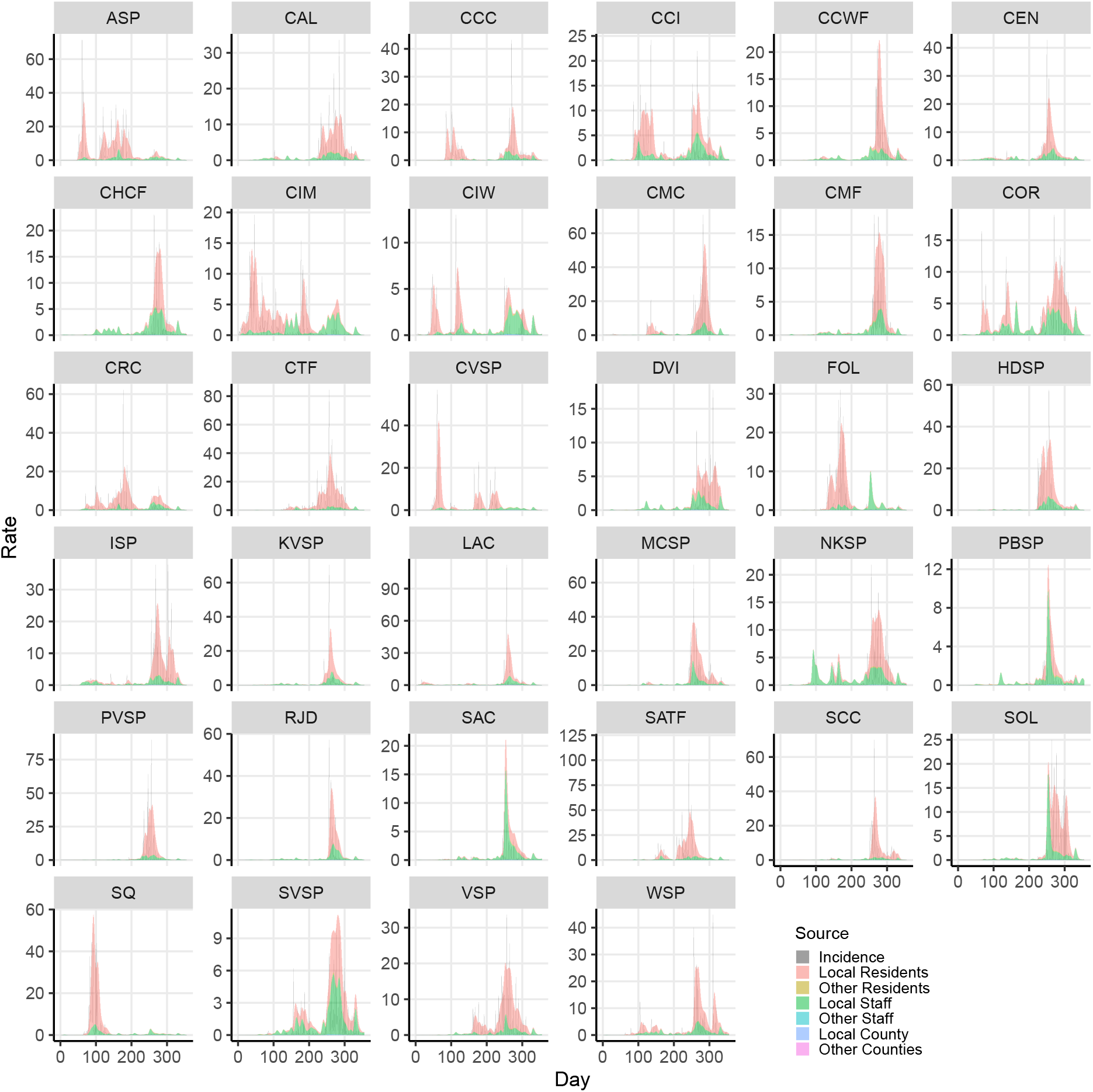
Incidence rates (gray) and intensities (colors, stacked) for resident cases by facility, in the maximum likelihood estimate for the well-mixed prisons nested model, with staff and resident exposures assumed identical.

**Figure B.6:**
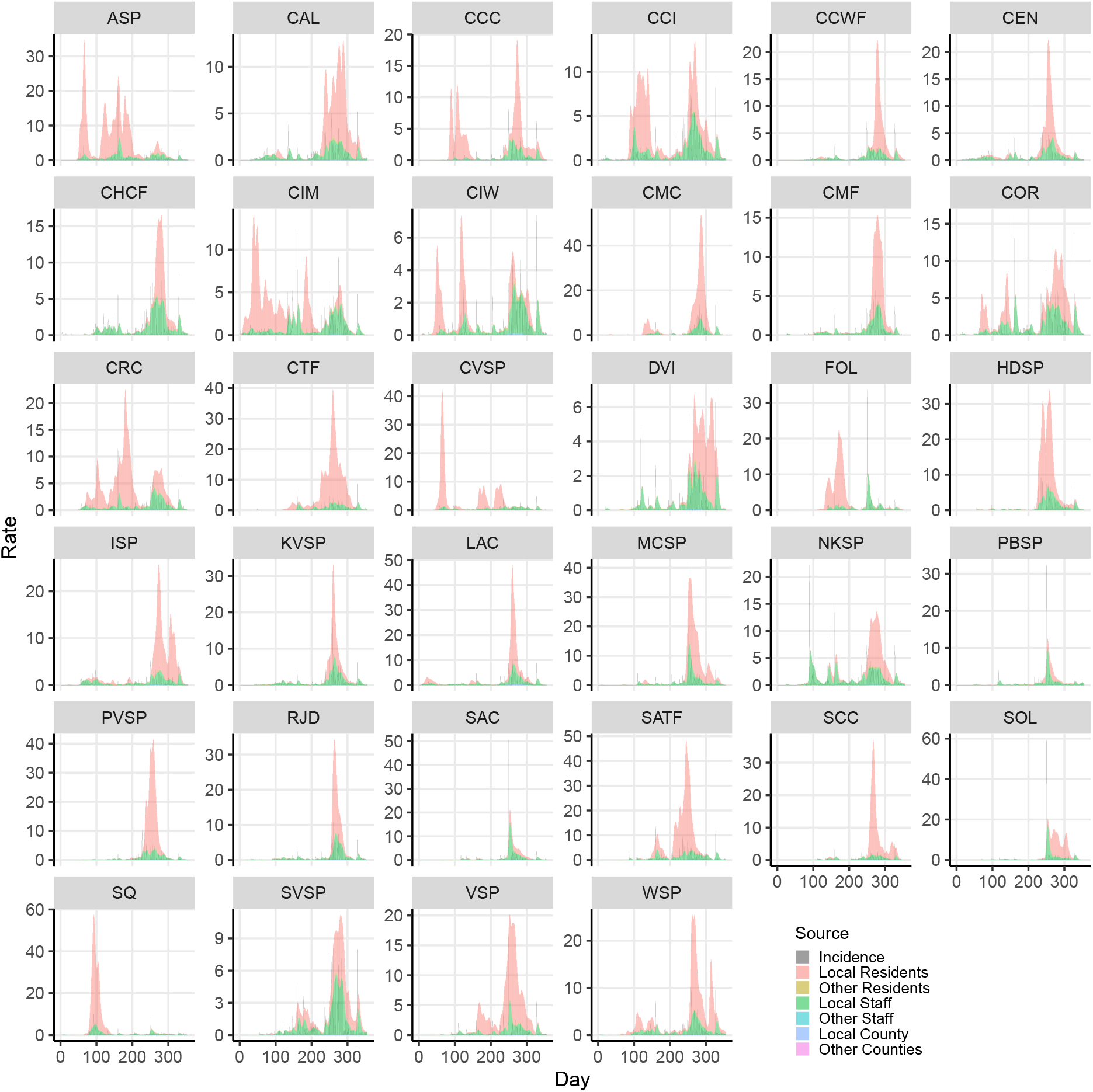
Incidence rates (gray) and intensities (colors, stacked) for staff cases by facility, in the maximum likelihood estimate for the well-mixed prisons nested model, with staff and resident exposures assumed identical.

**Table C.1:**
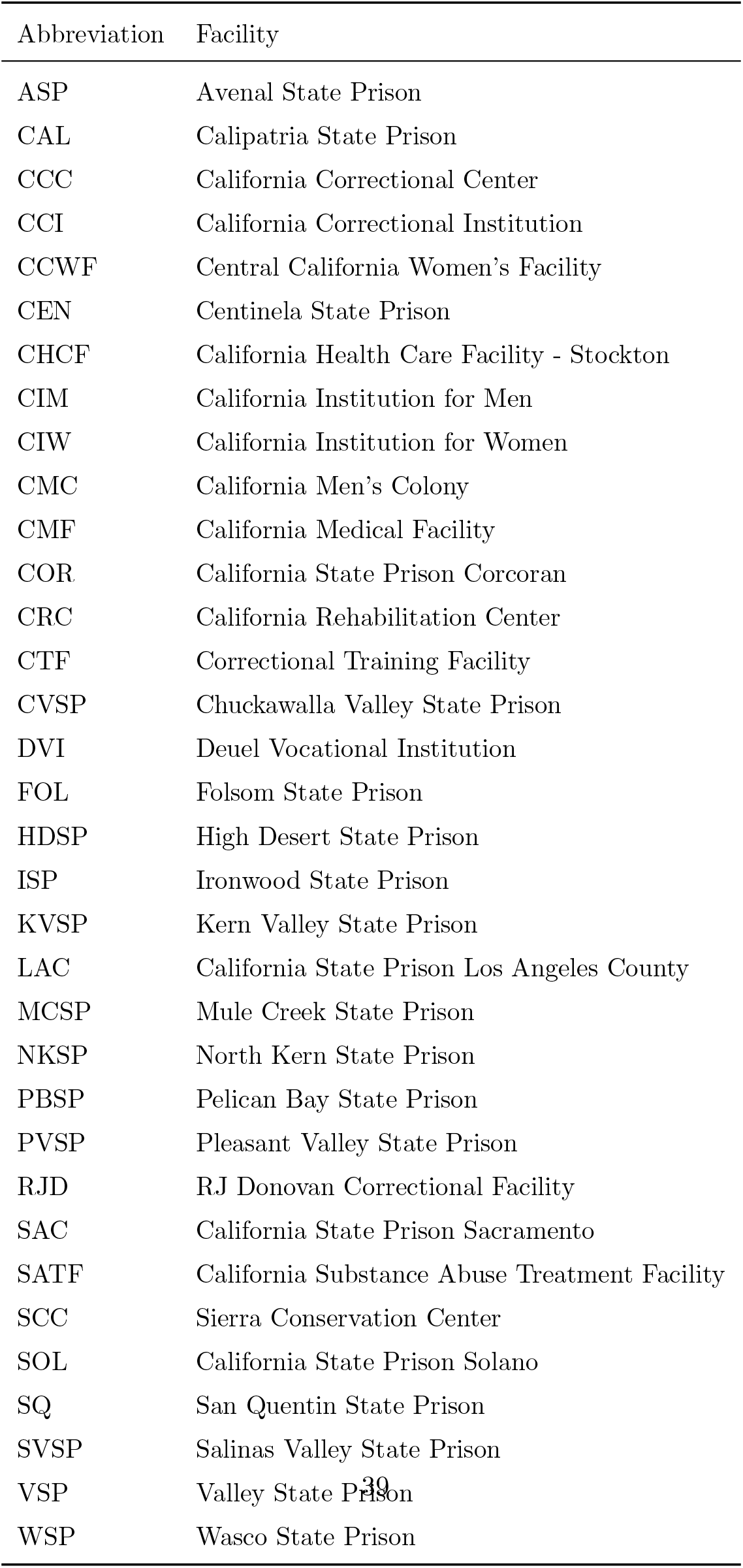
Abbreviated names of CDCR facilities.

| Abbreviation | Facility |
| --- | --- |
| ASP | Avenal State Prison |
| CAL | Calipatria State Prison |
| CCC | California Correctional Center |
| CCI | California Correctional Institution |
| CCWF | Central California Women's Facility |
| CEN | Centinela State Prison |
| CHCF | California Health Care Facility - Stockton |
| CIM | California Institution for Men |
| CIW | California Institution for Women |
| CMC | California Men's Colony |
| CMF | California Medical Facility |
| COR | California State Prison Corcoran |
| CRC | California Rehabilitation Center |
| CTF | Correctional Training Facility |
| CVSP | Chuckawalla Valley State Prison |
| DVI | Deuel Vocational Institution |
| FOL | Folsom State Prison |
| HDSP | High Desert State Prison |
| ISP | Ironwood State Prison |
| KVSP | Kern Valley State Prison |
| LAC | California State Prison Los Angeles County |
| MCSP | Mule Creek State Prison |
| NKSP | North Kern State Prison |
| PBSP | Pelican Bay State Prison |
| PVSP | Pleasant Valley State Prison |
| RJD | RJ Donovan Correctional Facility |
| SAC | California State Prison Sacramento |
| SATF | California Substance Abuse Treatment Facility |
| SCC | Sierra Conservation Center |
| SOL | California State Prison Solano |
| SQ | San Quentin State Prison |
| SVSP | Salinas Valley State Prison |
| VSP | Valley State Prison |
| WSP | Wasco State Prison |

## Notes

### Competing Interest Statement

Lee Worden reports a relationship with Office of the Federal Receiver for CCHCS that includes: funding grants.

### Author Declarations

https://github.com/uclalawcovid19behindbars/data/raw/master/historical-data/historical_facility_counts.csv https://data.chhs.ca.gov/dataset/f333528b-4d38-4814-bebb-12db1f10f535/resource/046cdd2b-31e5-4d34-9ed3-b48cdbc4be7a/download/covid19cases_test.csv

